# Dosimetric Impact of Residual Intrafraction Motion in Gated MR-Guided Prostate SBRT With Focal Dose Intensification

**DOI:** 10.64898/2026.09.11.26362864

**Authors:** Ka Ho Tam, Sandeep Jogi, Angela Sobremonte, Jared D. Ohrt, Dong Joo Rhee, Jinzhong Yang, Yao Ding, Kristy K. Brock, Peter A. Balter, Comron J. Hassanzadeh, Michael K. Rooney, Seungtaek Choi, Ryan Jin-hyung Park, Chad Tang, Krishnan R. Patel, Steven J. Frank, Phuoc T. Tran, Daniel E. Hyer, Neelam Tyagi, Ergys D. Subashi

## Abstract

**Purpose:** To quantify the dosimetric effect of residual intrafraction motion during gated MR-guided prostate stereotactic body radiotherapy with simultaneous integrated boost to the dominant intraprostatic lesion, and to derive and evaluate target-specific, population-based margins designed to mitigate motion-induced coverage loss.

**Materials and Methods:** Thirty patients were treated on a 1.5-T MR-Linac with 36.25, 40, and 45 Gy prescribed to the planning target volume, clinical target volume (CTV), and gross tumor volume (GTV), in 5 fractions. Across 150 fractions, segment-level dose matrices synchronized with time-resolved target displacements were combined to reconstruct per-fraction and cumulative dose. Directional prescription-isodose deviations were used to derive internal target volume margins, which were evaluated through offline replanning, gating-envelope analysis, and dose reconstruction.

**Results:** Median 95th-percentile motion ranges were 0.5, 1.4, and 2.9 mm in the left-right, anterior-posterior, and superior-inferior directions; these exceeded 3 mm in 0.7%, 13.2%, and 47.7% of fractions, respectively. The median treatment time was 13.2 min (IQR, 12.0-15.1 min) with a median gating duty cycle of 93.2% (80.6%-97.8%). Per-fraction median GTV deviations were −1.2% (−2.4% to −0.1%) for D95 and −5.6% (−15.3% to −0.2%) for V45Gy; corresponding CTV deviations were −0.5% (−1.1% to −0.1%) and −2.1% (−4.1% to −0.4%). Cohort-derived margins of 2/2/1/2/1/2 mm for the GTV and 1/1/1/2/1/2 mm for the CTV in the left/right/anterior/posterior/superior/inferior directions achieved sufficient coverage in approximately 94% of fractions in each direction. Gated delivery using the proposed margins improved coverage but may reduce duty cycle by an average of 6.9 ± 4.7 %.

**Conclusion:** Residual intrafraction motion produced directionally asymmetric coverage loss, with the largest effect on the boosted GTV. Motion-inclusive dose reconstruction enabled derivation of practical asymmetric margins that improved robustness while identifying the tradeoff between target coverage and delivery efficiency.

## INTRODUCTION

Stereotactic body radiotherapy (SBRT) is increasingly used for the definitive treatment of localized prostate cancer because it enables delivery of ablative doses in a short treatment course while maintaining acceptable toxicity. The clinical rationale for extreme hypofractionation is strengthened by randomized data supporting the safety of five-fraction prostate SBRT, as well as by the radiobiologic sensitivity of prostate cancer to larger fraction sizes (1). In parallel, focal dose escalation to the dominant intraprostatic lesion (DIL) has emerged as a strategy to improve tumor control without uniformly increasing dose to the whole gland. The FLAME trial demonstrated improved biochemical disease-free survival with focal boosting of the MRI-visible intraprostatic lesion while maintaining comparable toxicity, supporting the concept that dose intensification can be clinically beneficial when organ-at-risk (OAR) constraints are respected (2). These developments create a strong motivation for highly precise treatment delivery, particularly when high fractional doses and focal boosts are combined. While magnetic resonance-guided SBRT (MRgSBRT) has enabled margin reduction for whole-gland prostate treatment, focal DIL boosting introduces the additional challenge of treating a much smaller target embedded within a steep dose gradient. The DIL may therefore be more susceptible to residual intrafraction motion than the surrounding whole-prostate target. Whether currently employed motion-management strategies adequately preserve DIL dose coverage remains incompletely understood.

MRgSBRT provides several capabilities that are well suited to this setting and may mitigate some limitations of conventional SBRT. Superior soft-tissue contrast enables direct visualization of the prostate, DIL, and adjacent organs at risk, while online adaptive planning allows the treatment plan to be updated for the anatomy of the day (3, 4). These features have enabled reduction of planning margins in prostate SBRT (5). In the MIRAGE randomized trial, MRI-guided prostate SBRT using reduced margins was associated with significantly lower acute genitourinary and gastrointestinal toxicity compared with CT-guided SBRT using larger margins (6). However, online adaptation primarily addresses interfractional changes and pre-delivery anatomy. During a prolonged on-table interval required for online adaptive workflows, residual intrafraction motion can still occur during contour adaptation, plan optimization, verification imaging, and beam delivery. This residual motion becomes increasingly relevant as margins are reduced and dose gradients are intentionally sharpened around the prostate and organs-at-risk.

Intrafraction prostate motion has been characterized using implanted fiducials, electromagnetic transponders, ultrasound, and cine MRI. These studies have shown that prostate motion is time-dependent and anisotropic, with larger motion generally observed in the anterior-posterior and superior-inferior directions (7–9). In MR-guided treatment, continuous or repeated cine MRI provides a noninvasive method for direct soft-tissue motion monitoring without implanted surrogates or additional ionizing imaging dose (10, 11). Recent studies have demonstrated the feasibility of cine MRI-based prostate tracking, virtual couch shift correction, and real-time gating (12, 13). Nevertheless, displacement alone is an incomplete endpoint. The clinical relevance of a given motion trace depends on when the motion occurs during beam delivery, which segments are being delivered, the local dose gradient, the structure of interest, and whether deviations persist or are averaged out over the treatment course.

Dose reconstruction provides a more direct method for evaluating the consequence of intrafraction motion. By combining treatment delivery logs, segment-level dose information, and time-resolved target position, the delivered dose can be estimated and compared with the planned dose. Prior MR-Linac studies have demonstrated the feasibility of reconstructing delivered prostate dose from online MR imaging and machine log files, and have shown that intrafraction motion can produce fraction-specific deviations in target and organ-at-risk dose metrics (14, 15). More recent work has extended this concept toward online or real-time dose reconstruction, emphasizing that motion monitoring should ultimately be linked to delivered-dose assessment rather than geometric thresholds alone (16, 17). This is particularly important for DIL-boosted prostate SBRT, where small spatial deviations may have disproportionate effects on focal target coverage (18).

Real-time gating and baseline-shift correction can mitigate intrafraction motion but may interrupt or prolong delivery (12, 13, 19). Moreover, gating strategies are commonly defined relative to a predetermined structure, while residual dose errors may be directionally asymmetric and dependent on target size, location, and shape. A cohort-informed and target-specific planning volume may therefore better balance coverage and delivery efficiency.

Although prior studies have characterized prostate intrafraction motion and demonstrated the feasibility of motion-inclusive dose reconstruction, to our knowledge no prior study has quantified the dosimetric effect of residual intrafraction motion specifically within a gated MR-guided prostate SBRT workflow with focal DIL boosting. It therefore remains unclear whether geometric motion measurements alone adequately capture the dosimetric consequences of motion in this setting or how these observations should inform clinically implementable planning margins. We therefore sought to determine whether residual intrafraction motion disproportionately compromises focal boost coverage during gated MRgSBRT and whether motion-inclusive dose reconstruction could identify target-specific asymmetric margins that better preserve DIL coverage while maintaining treatment efficiency.

## MATERIALS AND METHODS

### Treatment planning and segment-level dose extraction

This retrospective study included 30 patients with localized prostate adenocarcinoma and at least one MRI-visible DIL treated with MRgSBRT on the 1.5-T Elekta Unity MR-Linac. All patients received 5 fractions, yielding 150 fractions for analysis. The prescription consisted of SIB dose levels of 36.25 Gy, 40 Gy, and 45 Gy to the planning target volume (PTV), clinical target volume (CTV) comprised of prostate gland and proximal seminal vesicles, and gross tumor volume (GTV/DIL), respectively. The CTV-to-PTV expansion was 3-4 mm isotropically and 2-3 mm posteriorly, at physician discretion. No margin was applied to the GTV/PTV. Prior to simulation, patients received a saline enema, and bladder volume was confirmed to be at least 100-150 cc using ultrasound (BladderScan i10, Verathon, Bothell, WA, USA). Before each fraction, patients again received a saline enema, and bladder volume was confirmed to be within 20%-30% of the simulation volume. A rectal spacer was used in all patients.

Treatment was delivered using coplanar fixed-gantry step-and-shoot intensity-modulated radiotherapy. Daily online adaptation was performed using the clinical adapt-to-position (ATP) or adapt-to-shape (ATS) workflow. Seventeen patients were treated with ATP plans for all five fractions, while the remaining thirteen patients had at least one ATS fraction. In five patients, two independent DIL subvolumes were contoured and planned separately and were analyzed as independent GTV structures in the per-fraction analysis.

Treatment plans were calculated in Monaco v6.2.2.0 using a Monte Carlo dose engine with 1% statistical uncertainty and a 3×3×3 mm³ dose grid. Segment-level dose matrices were exported for each delivered plan. Structure contours were converted to binary masks on the dose grid, with sampling coordinates generated at 1×1×1 mm³ spacing for subvoxel dose interpolation.

### Intrafraction motion monitoring and gating

Intrafraction motion was monitored using orthogonal sagittal and coronal balanced-contrast cine MRI acquired at 2.5 Hz per plane (20). The comprehensive motion management (CMM) system recorded three-dimensional CTV displacements relative to the setup position in synchrony with delivery log files (21, 22).

The beam was paused when the displaced CTV failed the prescribed volume-overlap criterion relative to the PTV. The threshold was initialized at 100% and could be reduced, typically to no less than 95%, to maintain delivery. When further reduction was required, a baseline-shift (BLS) plan re-centered the plan isocenter on the current target position and morphed the remaining segment apertures (12, 20).

For motion characterization, displacement traces were analyzed along the left-right (LR), anterior-posterior (AP), and superior-inferior (SI) directions. Displacements were measured relative to the target position on the daily setup MRI. Motion range was quantified using the 95th-percentile spread of the displacement distribution. To separate baseline drift from faster motion components, each displacement trace was decomposed using a 20-second moving-average low-pass filter and a high-pass filter with a 0.05 Hz cutoff (see Supplementary Figure S1 for illustration). The probability of displacement exceeding 1, 2, and 3 mm thresholds was evaluated as a function of time for each cardinal direction. The fraction of time spent outside each threshold during treatment was also calculated.

### Dose reconstruction

Dose reconstruction was performed by combining segment-level dose matrices, machine delivery logs, and time-resolved target displacement. Motion was modeled as rigid translation of the target relative to the static dose distribution. Under this approximation, a target displacement vector *δ*(*t*) at time *t* is equivalent to sampling the segment-dose matrix at shifted structure coordinates *r* + *δ*(*t*), where *r* denotes the structure sampling coordinate.

For each structure coordinate, the reconstructed dose was computed by summing the monitor-unit-weighted dose contribution from all beam-on time points:

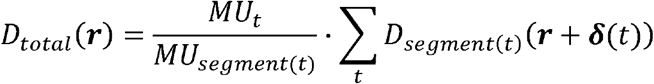

where *D_segment_*_(*t*)_ is the segment-level dose matrix corresponding to the active beam segment at time *t*, *δ*(*t*), is the measured target displacement vector, *MU_t_* is the monitor units delivered during the time interval, and *MU_segment_*_(*t*)_ is the total monitor units planned for the corresponding segment. Dose values at shifted sampling coordinates were computed using trilinear interpolation from the surrounding dose-grid voxels.

For fractions in which a BLS plan was delivered, reconstruction used the segment-level dose matrices corresponding to the plan that was active at each time point. Thus, dose delivered before the correction was reconstructed using the original adapted plan, while dose delivered after the correction was reconstructed using the BLS plan.

Reconstructions performed with zero displacement were used as a quality assurance step to confirm that the pipeline reproduced the planned dose-volume histogram.

Planned and reconstructed dose-volume histogram (DVH) metrics were calculated for each structure at each fraction. Dosimetric deviations were reported as normalized differences:

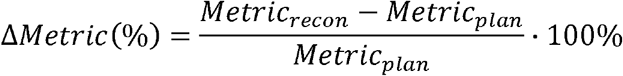

Target coverage metrics included D95% and V45Gy for the GTV and D95% and V40Gy for the CTV. Near-maximum dose was evaluated using D0.03cc. Each delivered fraction was analyzed as a separate observation. Per-patient cumulative dose was estimated by summing the five per-fraction reconstructed dose distributions for each patient. For patients treated entirely using ATP structure sets, dose was accumulated directly in the CT-based reference frame. For patients with ATS fractions, target structures were rigidly transferred to the reference CT frame before dose accumulation to mitigate uncertainties due to deformable image registration. A schematic diagram of the workflow is presented in Figure 1.

**Figure 1.**
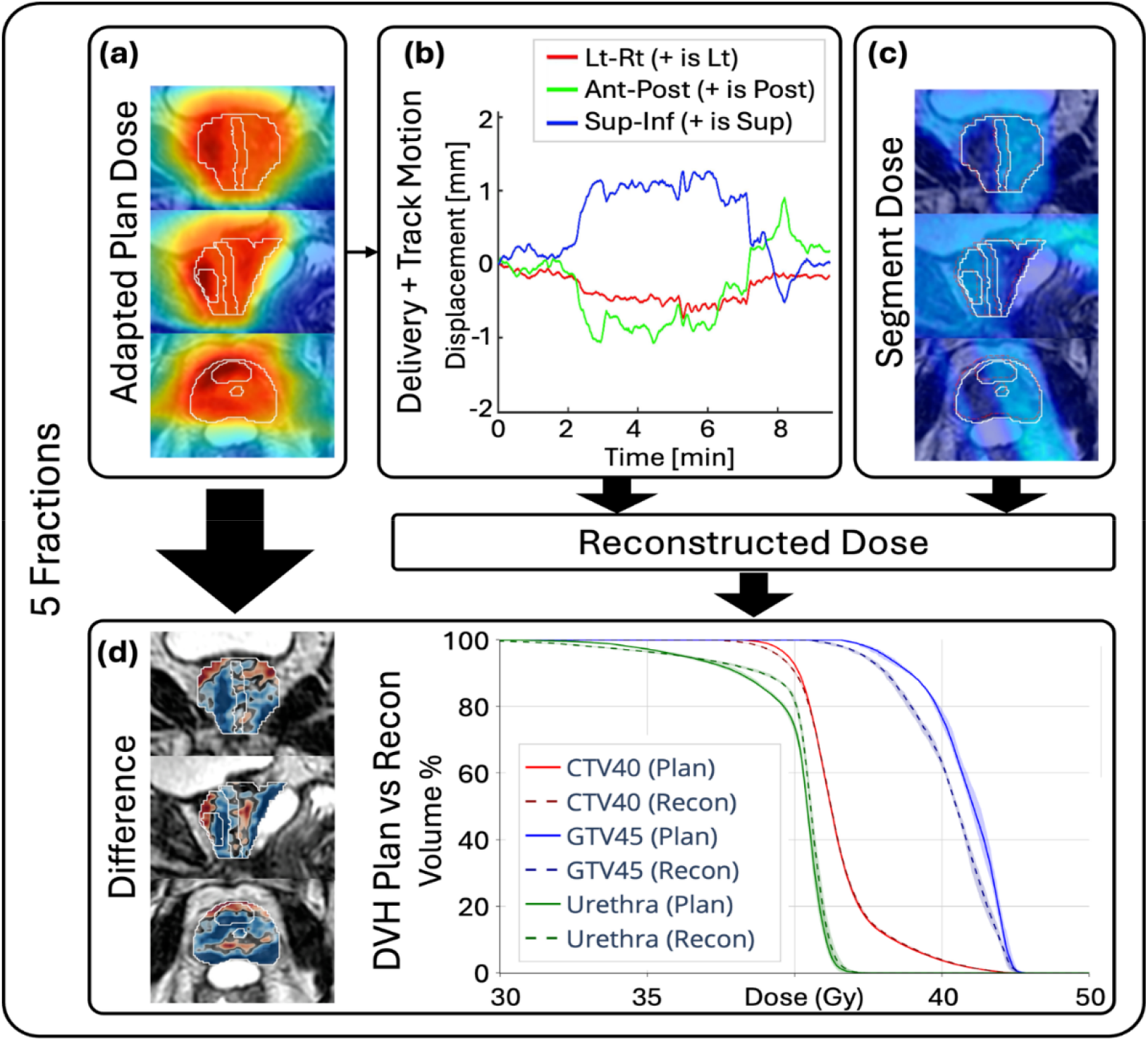
Method overview for dose reconstruction. (a) For each fraction, segment-level dose matrices from the adapted treatment plan were exported from the treatment planning system and used as input to the reconstruction pipeline. (b) During treatment, target displacement vectors were recorded in synchrony with delivery log files. (c) Dose was reconstructed by accumulating the segment-level dose contributions according to the delivered monitor units and measured target position at each time point. (d) Reconstructed dose distributions were compared with the planned dose distributions using spatial dose-difference maps and DVH metrics for each fraction and across the treatment course.

The evaluated metrics were ΔV40Gy for the CTV, ΔV45Gy for the GTV, ΔD95 and ΔD0.03cc for both targets.

### Spatial analysis of prescription isodose-line deviations

Underdosed CTV or GTV voxels outside the corresponding prescription isodose line (IDL) were identified in the planned and reconstructed dose distributions. The distance between each underdosed voxel and the prescription IDL was measured along the cardinal directions. The directional IDL-to-target distance was defined as the 95th-percentile distance, with 0 mm assigned when no underdosed voxels were present. Directional deviation was calculated as the reconstructed minus planned distance.

For each target and direction, the expansion required to encompass deviations in 90% of fractions was calculated and rounded up to the nearest millimeter. The resulting CTV expansion defined the cohort-informed planning internal target volume (pITV). Planning feasibility was evaluated by reoptimizing reference plans using the proposed CTV and GTV expansions. The dosimetric benefit was further evaluated in 10% of fractions with the largest V45Gy deviations by regenerating adapted plans offline and repeating motion-inclusive dose reconstruction. The number of fractions in which patient-specific margins exceeded the proposed population margins were also recorded.

The relationship between residual target motion and reconstructed dose deviations was evaluated using per-fraction mean displacement. Associations between mean displacement and changes in D95, prescription-dose volume coverage, and D0.03cc were analyzed separately for the CTV and GTV. Linear mixed-effects regression modeling was used to account for variability across fractions within patients. Structure volume was evaluated as an additional predictor to assess whether volume modified the relationship between displacement and dosimetric deviation.

Geometric robustness was evaluated using gating-envelope coverage (GEC) structures generated by expanding the CTV isotropically by 0–3 mm. For each recorded CMM displacement, the percentage of CTV contained within each GEC was calculated. The clinical PTV-based GEC was compared with the cohort-informed pITV. The probability that 95%, 98%, or 100% of the CTV remained within each structure was evaluated as a function of time after beam-on. CMM duty cycle was defined as the percentage of time for which CTV containment satisfied the specified gating threshold. Non-CMM duty cycle was also calculated as the percentage of time during which the treatment beam was not interrupted due to MLC or gantry movements and internal machine safety checks. To account for overlaps between these events and include beam stops due to unplanned machine events, the total duty cycle was calculated as the percentage of time during which the treatment beam was on.

### Statistical analysis

Continuous variables were summarized using the median and interquartile range unless otherwise specified. Differences among fractions were assessed using the Kruskal– Wallis test. Associations between mean displacement and dosimetric outcomes were evaluated using linear mixed-effects models with a patient-specific random intercept.

Confidence intervals were estimated using patient-level bootstrapping. Statistical analyses were performed using Python v3.12, with a two-sided *p* < .05 considered statistically significant.

## RESULTS

### Treatment delivery and intrafraction motion

Dose reconstruction was completed for all 150 fractions. The median treatment time was 13.2 min (IQR, 12.0–15.1 min), while median CMM, non-CMM, and total duty cycles were 93.2%, 57.0%, and 45.0%, as shown in Table 1. Thirty-two fractions (21%) in 16 patients included at least one mid-delivery BLS plan. This yielded 175 per-fraction GTV observations.

**Table 1.** Summary of metrics characterizing intrafraction motion and duty cycle across all 150 fractions. Values are reported as median (IQR); Prob(p95<3 mm) is reported as the percentage of fractions satisfying the specified condition. Target drift is characterized by LP-filtered measurements, whereas higher-frequency components (eg. cardiac, respiratory, noise) motion is characterized by HP-filtered measurements. Drift was the dominant component of target motion and beam stops due to CMM were comparatively much smaller than those from non-CMM events. p95 = 95^th^-percentile spread of target displacement; LP = low-pass filtered (20-sec moving average); HP = high-pass filtered (>0.05 Hz); CMM = comprehensive motion management.

|  | LR | AP | SI |
| --- | --- | --- | --- |
| **p95 [mm]** | 0.5 (0.7) | 1.4 (1.3) | 2.9 (2.8) |
| **Prob (p95 < 3 mm) [%]** | 99.3 | 86.8 | 52.3 |
| **p95 LP [mm]** | 0.3 (0.4) | 1.3 (1.2) | 2.0 (2.4) |
| **p95 HP [mm]** | 0.2 (0.3) | 0.5 (0.3) | 1.0 (0.6) |
| **CMM Duty Cycle [%]** | 93.2 (16.9) |  |  |
| **Non-CMM Duty Cycle [%]** | 57.0 (11.0) |  |  |
| **Total Duty Cycle [%]** | 45.0 (9.7) |  |  |

Mean displacement from setup position increased with elapsed treatment time and was directionally anisotropic, with the largest excursions occurring in the inferior and posterior directions, summarized in Figure 2. The median (IQR) p95 motion ranges were 2.9 (2.8) mm in the SI direction, 1.4 (1.3) mm in the AP direction, and 0.5 (0.7) mm in the LR direction. The p95 range exceeded 3 mm in 47.7%, 13.2%, and 0.7% of fractions in the SI, AP, and LR directions, respectively. Motion decomposition showed that slow drift was the dominant component of SI motion: the median p95 SI range was 2.0 mm for the low-pass component and 1.0 mm for the high-pass component. The corresponding low-pass and high-pass ranges were smaller in the AP and LR directions. Across 150 fractions, the correlation between the SI and AP displacement was significant (LME, p<<0.05). The correlation between LR and AP or LR and SI displacement was not significant (LME, p>0.05).

**Figure 2.**
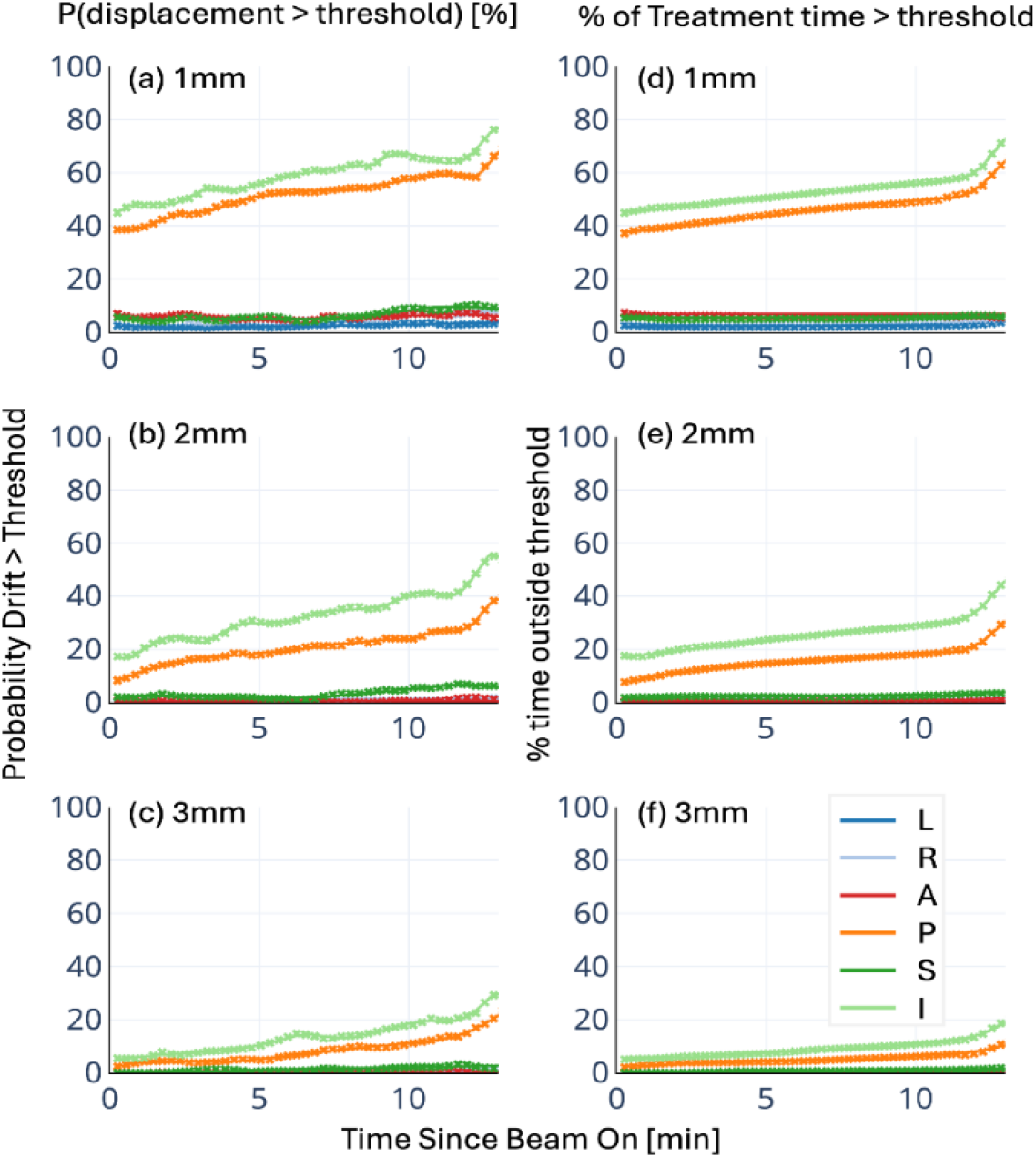
Characterization of intrafraction motion across 150 fractions, 30 patients. (a–c) Probability that target drift exceeds the displacement threshold (1mm, 2mm, or 3mm) at a given time, resolved along the cardinal directions. (d– f) Fraction of treatment time spent outside the displacement threshold. Time zero denotes beam-on; displacement was measured relative to the target position on the daily setup MRI. Intrafraction drift increased with elapsed treatment time and was directionally anisotropic, with the largest excursions observed in the inferior and posterior directions. Minimal drift was observed along the left, right, anterior, and superior directions. L/R = left/right; A/P = anterior/posterior; S/I = superior/inferior.

Beam-on and continuously measured motion distributions could differ because of gating and delivery timing (Supplementary Figure S1). Frequency analysis identified lower-frequency respiratory-related components, while the dominant 2.5-Hz SI component corresponded to the imaging frequency and was attributed to measurement noise (Supplementary Figure S2).

### Dosimetric effect of intrafraction motion

Residual motion preferentially compromised focal SIB coverage, with dosimetric coverage losses approximately 2-3-fold greater for the boosted GTV than for the whole-prostate CTV, as shown in Figure 3 and Supplementary Table S1. Representative outlier traces were selected based on the lowest minimum time-dependent D95 values to illustrate variability. Median per-fraction GTV deviations were −1.2% (IQR, −2.4% to −0.1%) for D95 and −5.6% (−15.3% to −0.2%) for V45Gy; corresponding CTV deviations were −0.5% (−1.1% to −0.1%) for D95 and −2.1% (−4.1% to −0.4%) for V40Gy. Cumulative deviations were smaller but remained negative: −0.8% and −4.0% for GTV D95 and V45Gy and −0.3% and −1.2% for CTV D95 and V40Gy. Deviations did not differ significantly among fractions (*P* > .05). Supplementary Figure S3 presents the results of the quality assurance step on the computation pipeline comparing reconstructions performed with zero displacement to confirm that the pipeline reproduced the planned dose-volume histogram.

**Figure 3.**
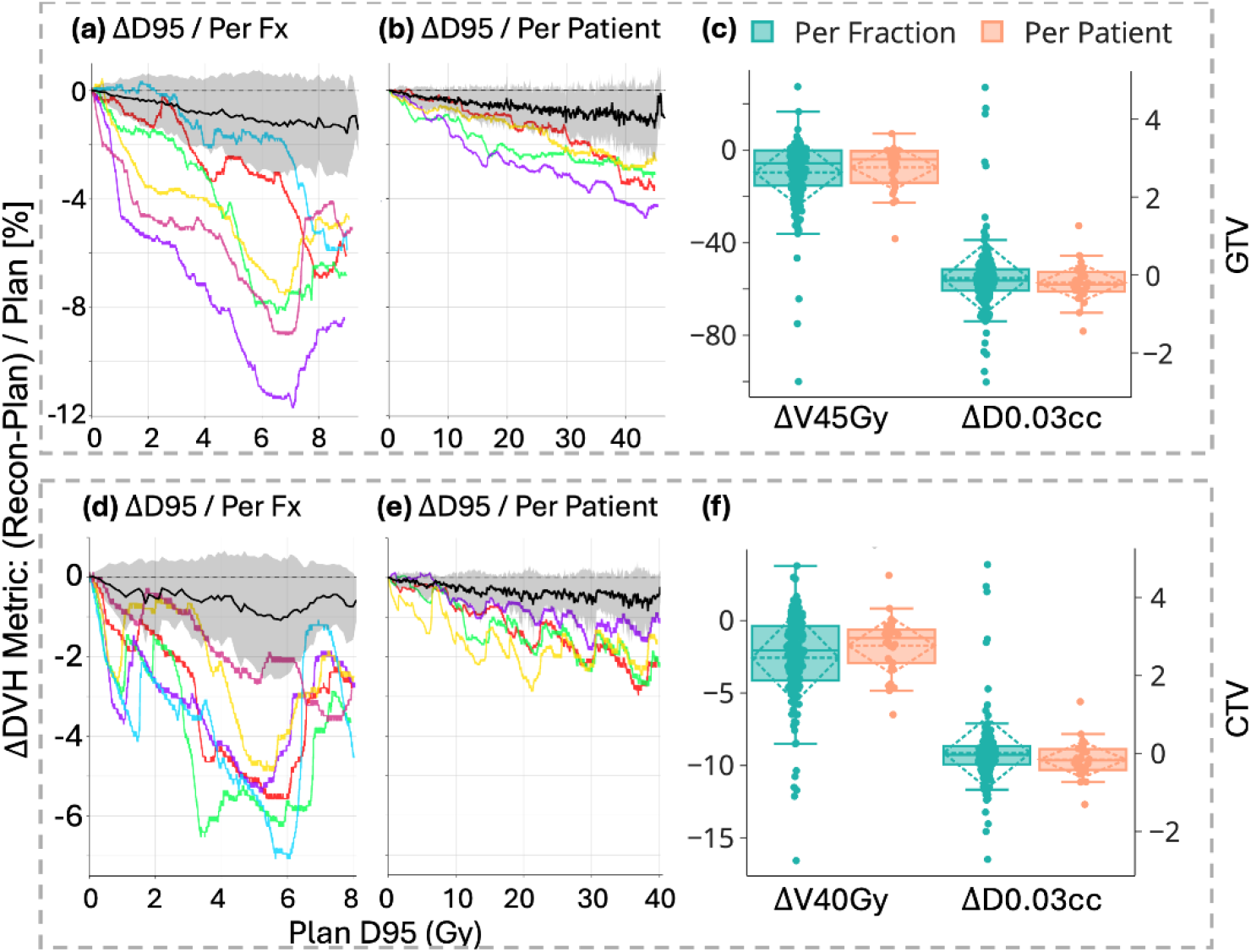
Dosimetric deviations, calculated as ΔMetric = (Recon − Plan)/Plan×100%, for the GTV45 (a–c) and CTV40 (d–f). Panels (a,d) show per-fraction changes in D95 as a function of planned D95, and panels (b,e) show the corresponding cumulative per-patient changes. Colored lines denote outlier fractions or patients, while the black line and shaded region denote the cohort mean ± standard deviation. Panels (c,f) summarize per-fraction and per-patient distributions of prescription-dose coverage (ΔV45Gy for the GTV and ΔV40Gy for the CTV; left axes) and near-maximum dose (ΔD0.03cc; right axes) deviations. Negative values indicate reduced reconstructed dose relative to the planned dose. Coverage losses were larger per fraction than per patient, indicating partial averaging over the treatment course, whereas ΔD0.03cc remained small and approximately symmetric about zero. D95 = dose received by 95% of the target volume; V40Gy/V45Gy = percentage of the CTV/GTV receiving at least 40 Gy/45 Gy, respectively; D0.03cc = dose to the hottest 0.03 cc.

Mean intrafraction displacement was significantly associated with reductions in D95 and prescription-dose volume coverage for both the CTV and GTV, as demonstrated in Figure S4. In contrast, ΔD0.03cc was not significantly associated with mean displacement. Structure volume was not a significant predictor of coverage loss but was positively associated with GTV ΔD0.03cc; no corresponding association was observed for CTV ΔD0.03cc.

### Spatial coverage deviations and margin adaptation

IDL-to-target deviations were anisotropic, with the largest coverage losses inferiorly and posteriorly, as shown in Figure 4(a,b). In each cardinal direction, expansions required to encompass deviations in 90% of fractions were 1.2/1.4/0.8/1.6/0.3/2.0 mm for the GTV and 1.0/0.7/0.9/1.3/0.4/1.8 mm for the CTV in L/R/A/P/S/I respectively. These were rounded to 2/2/1/2/1/2 mm and 1/1/1/2/1/2 mm, respectively, encompassing deviations in approximately 94% of fractions. All planning goals were achieved in the replanned cases.

**Figure 4.**
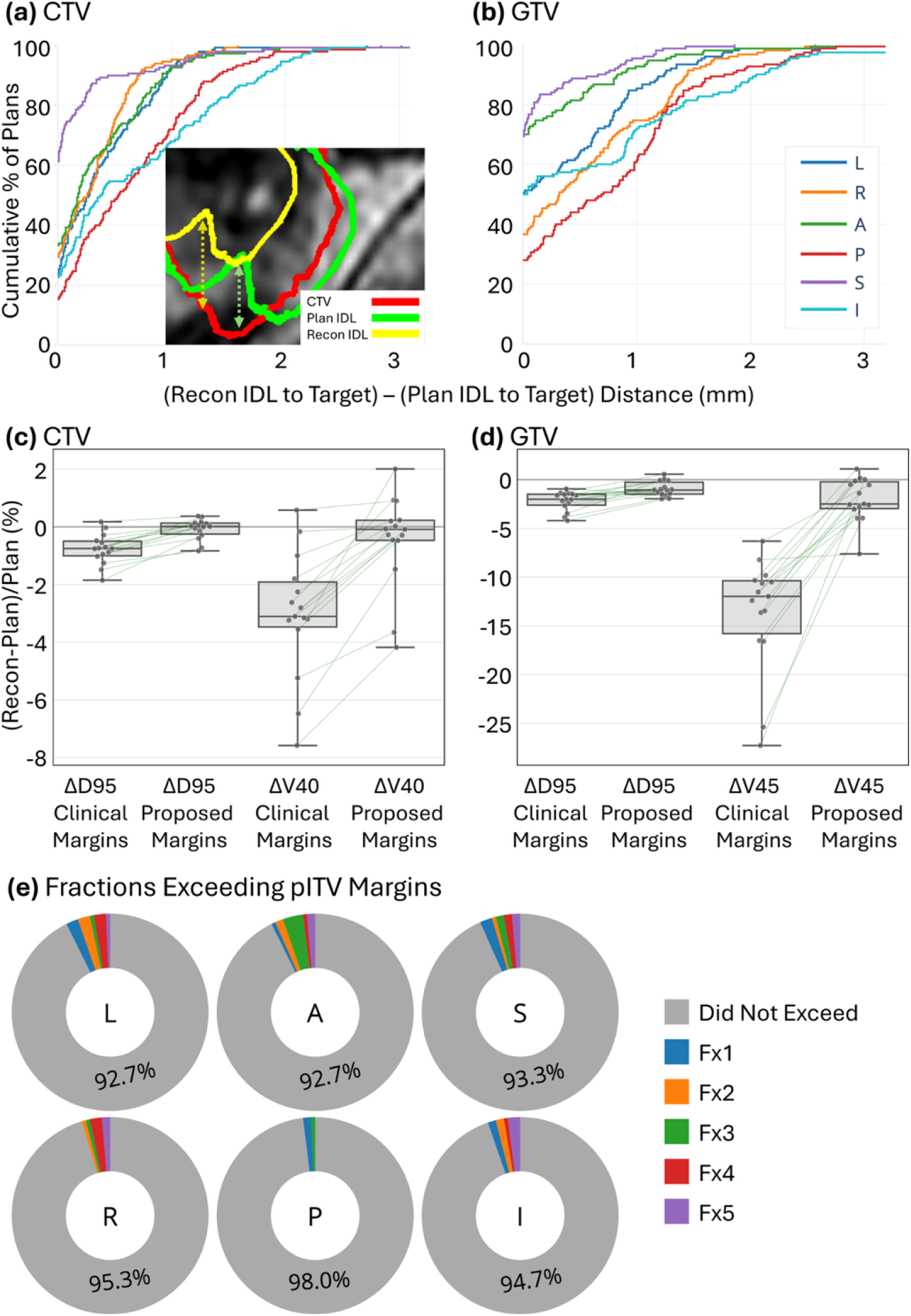
Prescription IDL deviations and dosimetric effect of margin expansion. Cumulative distribution of IDL-to-target distance deviations, calculated as the reconstructed prescription IDL-to-target distance minus the corresponding planned distance, are shown for the (a) CTV and (b) GTV, resolved along the cardinal directions. The inset illustrates the measurement on a representative slice: target contour in red, planned IDL in green, and reconstructed IDL in yellow. Smaller deviations indicate closer agreement between reconstructed and planned coverage. Deviations were anisotropic, with the largest values in the inferior and posterior directions. (c,d) Paired per-fraction deviations in target coverage metrics between plans using the clinical margins and plans using the proposed population margins. Dosimetric goals were satisfied in all reference plans recreated with population margins. Lines connect results from the same fractions, and boxplots summarize the cohort distributions. The proposed margins shifted coverage deviations toward zero, indicating improved agreement between reconstructed and planned dose. (e) The number of fractions, represented as percentage over all fractions, in which patient-specific margins exceeded the proposed population margins. L/R = left/right; A/P = anterior/posterior; S/I = superior/inferior.

In the 10% of fractions with the largest V45Gy deviations, the proposed margins shifted D95 and prescription-dose coverage deviations toward zero while maintaining clinical planning goals, summarized in Figure 4(c,d). Figure 4(e) shows the percentage of fractions in which the patient-specific margin exceeded the proposed population margin.

Figure 5(a-c) reveals that increasing the GEC margin improved CTV containment for a given displacement, while Figure 5(d,e) demonstrates the tradeoff between geometric sensitivity and delivery efficiency. Clinically, CTV coverage decreased with time and with more restrictive overlap criteria for both PTV- and pITV-based gating. The smaller pITV provided higher dose coverage and reduced duty cycle by an average of 6.9 ± 4.7%, calculated as the mean difference between duty cycles using PTV or pITV as GEC across gating thresholds of 90%-100%.

**Figure 5.**
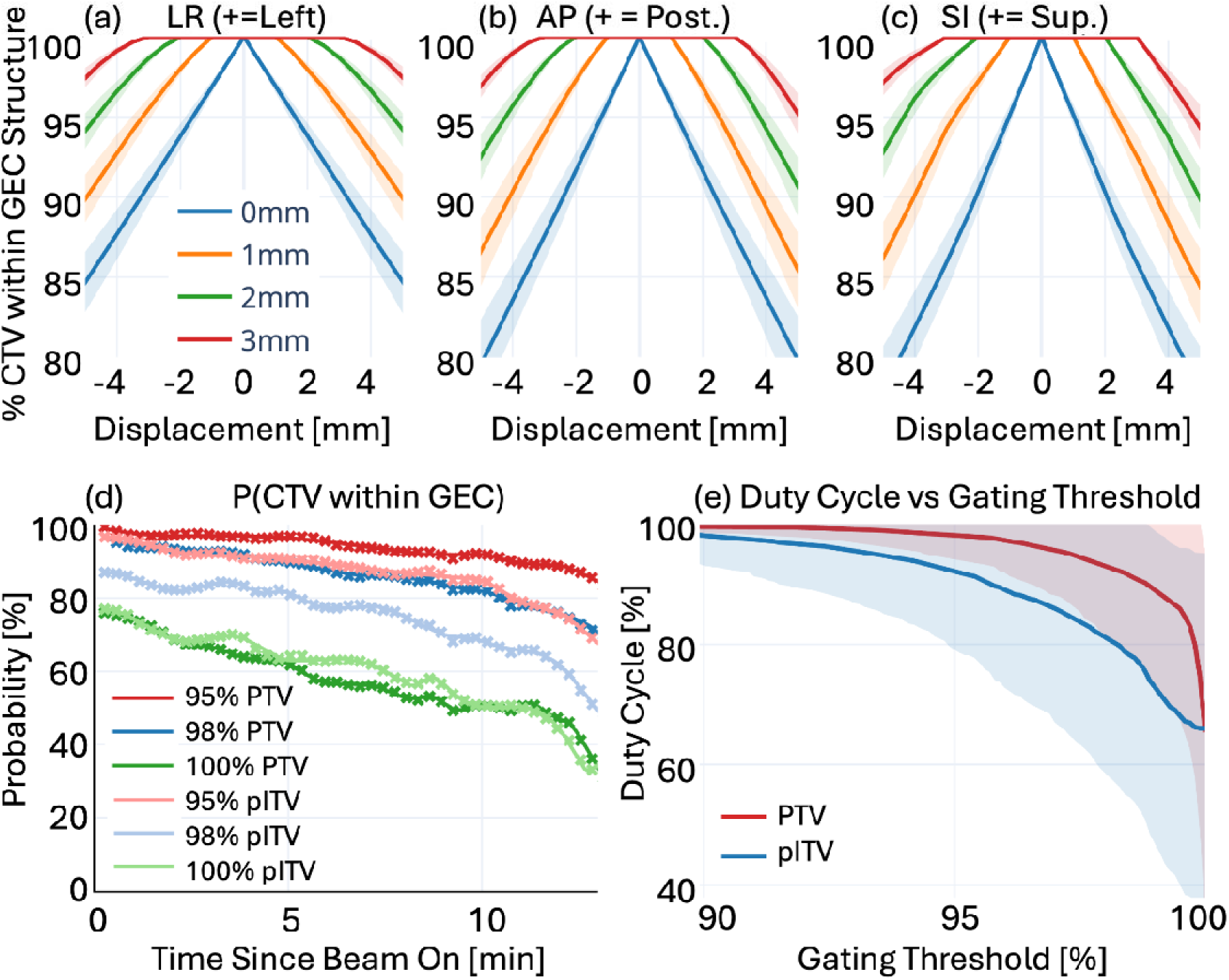
Sensitivity of CTV coverage to residual displacement and gating configuration, evaluated using intrafraction motion from 150 fractions, 30 patients. (a–c) Percent volume of CTV contained within the gating envelope coverage (GEC) structure as a function of target displacement, for GEC = CTV + 0, 1, 2, 3 mm margin; shaded bands denote cohort standard deviation. (d) Probability that the CTV remained within the GEC structure as a function of time since beam-on, evaluated at 95%, 98%, and 100% CTV coverage for the PTV- and pITV-based GEC structures. (e) Duty cycle as a function of gating threshold for the PTV and pITV structures; lines and shaded bands denote the cohort mean and standard deviation. Although margin expansion improved geometric robustness to displacement, differences between PTV- and pITV-based gating configurations were comparatively small when evaluated using the probability of CTV containment and estimated duty cycle. PTV = CTV expanded by the prescription margins; pITV = CTV expanded by 1 mm isotropically, with a 2-mm expansion in the inferior and posterior directions. pITV=planning internal target volume.

We summarize exploratory patient- and physician-reported GU/GI toxicity events in Supplementary Materials. Toxicity outcomes were favorable, with one grade 2 GU event among 26 evaluable patients and no acute GI toxicity. IPSS changes were variable but tended to be larger among patients with higher GU toxicity grades. Note that this study was not powered to determine whether reconstructed dose was associated with toxicity. Larger studies have reported associations between prolonged motion, motion direction, and urinary adverse events (9, 17), suggesting that cumulative motion-inclusive dose may warrant prospective evaluation as a predictor of clinical outcome.

## DISCUSSION

This study evaluated the effect of residual intrafraction motion on target coverage during gated MRgSBRT for prostate with SIB to the dominant intraprostatic lesion. The principal contribution of this work is the demonstration that motion-inclusive dose reconstruction identifies a dosimetric vulnerability for focal DIL boosting that is not fully captured by geometric motion alone and provides a framework for target-specific margin design. Median coverage losses were approximately 2–3-fold greater for the boosted GTV than for the CTV, indicating greater sensitivity of focal boost coverage to residual motion, and highlighting an important limitation of current motion-management paradigms that rely on geometric criteria. Coverage loss was spatially anisotropic and greatest in the inferior and posterior directions, consistent with the predominant direction of prostate drift. Based on these findings, we derived and evaluated population-based asymmetric margins intended to mitigate motion-induced coverage loss while maintaining an acceptable gating duty cycle.

Intrafraction motion increased with elapsed treatment time and was largest in the SI direction. The median p95 SI motion range was 2.9 mm, and the p95 range exceeded 3 mm in nearly half of the fractions. Signal decomposition showed that slow drift accounted for most of the SI motion, whereas the higher-frequency component was smaller. This behavior is consistent with previous cine-MRI and real-time tracking studies showing that prostate motion is time dependent, anisotropic, and frequently characterized by gradual drift (16–18). Supplementary Figure S1 also demonstrates that the continuously measured and beam-on motion distributions may differ. Thus, motion magnitude alone does not fully describe the motion relevant to dose delivery, because the dosimetric effect also depends on when the displacement occurs during the delivery of a given fraction and how steep the dose gradient is when the segment is being delivered.

Residual motion primarily affected coverage metrics, while changes in D0.03cc were small and approximately centered around zero. Per fraction, median GTV D95 and V45Gy deviations were −1.2% and −5.6%, respectively, compared with −0.5% and −2.1% for CTV D95 and V40Gy. The distributions were wider for the GTV and this greater sensitivity is expected because the DIL is small, is often surrounded by a steep dose gradient, and was treated without a dedicated margin. A displacement that remains within the CTV-to-PTV expansion may therefore still move part of the DIL outside the 45-Gy isodose. This is relevant because the benefit of focal boosting depends on reliably delivering the intended dose to the MRI-visible lesion while respecting OAR constraints. All OAR constraints were met in the evaluated reference replans using the expanded CTV and GTV margins.

Cumulative reconstruction showed that fraction-specific deviations partially averaged over the treatment course but did not eliminate the coverage bias. Median cumulative GTV deviations remained −0.8% for D95 and −4.0% for V45Gy. The corresponding CTV deviations were −0.3% and −1.2%. No systematic trend between DVH deviations and fraction number was observed, indicating that no specific fractions were more susceptible to motion. Instead, while margin exceedances were uncommon, these deviations occurred variably among treatment sessions. These findings support dose assessment across the full treatment course rather than relying on motion observed during a single fraction.

The magnitude of the CTV deviations was comparable with previous dose-reconstruction studies. Klucznik et al. reported a mean per-fraction CTV D95 deviation of approximately −0.5% with intrafraction image guidance and repositioning, while accumulated deviations approached zero (16). Previous cine-MRI studies similarly found modest average dose changes but larger deviations in individual fractions (23). Brennan et al. observed progressive reductions in DIL coverage between planning, verification, and post-treatment imaging during MR-guided focal-boost SBRT (18). The present study complements these observations by combining time-resolved target motion with segment-level dose matrices and delivered monitor units in a gated workflow. This preserves the temporal relationship between displacement and beam delivery, which is important in studying the interplay effect (24). Our analysis extends these observations by using motion-inclusive dose reconstruction to derive and evaluate clinically implementable asymmetric margins for both whole-gland and focal boost targets. Thus, we believe this work shifts emphasis from describing motion to informing treatment planning.

Importantly, our results suggest that conventional geometric motion statistics may be insufficient for margin design, as equivalent displacements can have substantially different dosimetric consequences depending on target size, boost location and treatment timing. The directional IDL analysis provided a dose-based framework for margin design. To remove IDL-to-target deviations in 90% of fractions, the required expansions were largest inferiorly and posteriorly. After rounding for clinical implementation, the proposed population-based expansions were 2/2/1/2/1/2 mm for the GTV and 1/1/1/2/1/2 mm for the CTV in the L/R/A/P/S/I directions. These margins removed the evaluated deviations in approximately 94% of fractions in each direction. In addition, all clinical planning goals were maintained when the expanded structures were used to reoptimize the evaluated reference plans, and dose reconstruction of the most affected fractions showed that the proposed margins shifted D95 and prescription-dose coverage deviations toward zero.

These expansions should be interpreted as internal planning margins intended to account specifically for residual intrafraction motion within an online-adaptive and gated workflow. They are not replacements for conventional CTV-to-PTV margins, which account for multiple sources of geometric uncertainty (25–28). Their asymmetric design reflects the observed inferior–posterior coverage pattern and avoids uniformly increasing the treated volume (29). Previous cine-MRI studies have supported CTV-to-PTV margins of approximately 3 mm for five-fraction MR-guided prostate SBRT (5, 30–33). The present analysis complements these studies by deriving dose-informed, target-specific expansions, particularly for the focal boost. Margins adequate for whole-prostate coverage may not fully protect the focal-boost target.

The same population-based asymmetric expansion was used to define the cohort-informed pITV for the gating analysis. Increasing the gating-envelope margin improved CTV containment for a given displacement. The larger PTV-based gating structure provided greater containment than the smaller pITV, whereas pITV-based gating produced a lower estimated duty cycle, particularly at thresholds approaching complete CTV containment. Small differences at a 95% containment criterion suggest that moderate overlap thresholds may retain much of the geometric benefit without the delivery penalty associated with requiring complete GEC coverage. More generally, the results illustrate that a smaller, dose-informed target expansion can improve sensitivity to motion while minimizing the effect on dose coverage. Selection of the gating structure and threshold should therefore balance target robustness against treatment efficiency (34).

This study has several limitations. Motion was modeled as rigid translation relative to a static dose distribution and therefore did not account for target deformation, differential motion of adjacent structures, or changes in radiological path length. CTV-derived displacement was applied to the GTV under the assumption that these structures moved together. The CMM tracking signal is also subject to image-resolution and position-estimation uncertainty (22). Finally, the proposed margins were derived and evaluated in a single-institution cohort, and the planning assessment was limited to a subset of cases. Prospective and external validation is therefore required before clinical implementation, including evaluation of their effects on organ-at-risk dose.

## CONCLUSIONS

This study demonstrated that motion-inclusive dose reconstruction can identify target-coverage effects from residual motion during gated focal-boost MRgSBRT that are not readily apparent from geometric motion measurements alone. This framework may provide a more clinically relevant basis for future target-specific margin design and motion management strategies. Although dosimetric deviations partially averaged over five fractions, a cumulative coverage loss remained at completion of treatment. Cohort-derived asymmetric expansions of 2/2/1/2/1/2 mm for the GTV and 1/1/1/2/1/2 mm for the CTV in the L/R/A/P/S/I directions mitigated coverage loss in 94% of fractions per direction while maintaining clinical planning goals. These findings support the use of motion-inclusive dose assessment to inform population-based margins and gating strategies that balance target robustness against delivery efficiency.

## Data Availability

The data underlying this study are not publicly available and are subject to institutional restrictions. Requests for access to data are subject to review and approval by The University of Texas MD Anderson Cancer Center and may require additional regulatory and data-use agreements.

