## Supplementary Materials for "Dosimetric Impact of Residual Intrafraction Motion in Gated MR-Guided Prostate SBRT With Focal Dose Intensification"

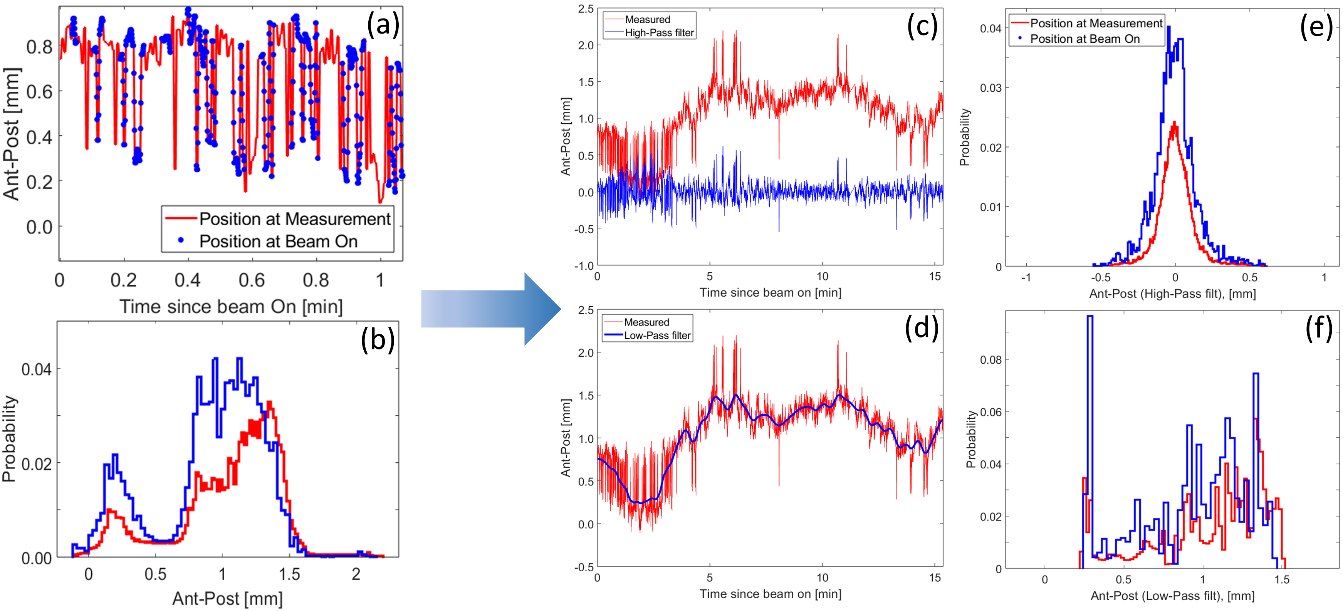

Figure S1. Representative decomposition of intrafraction target motion into high- and low-frequency components. (a) Time-dependent prostate target position in the anterior–posterior (AP) direction during the first minute after beam-on; red denotes all measured positions and blue markers denote positions sampled during beam-on. (b) Probability distributions of measured and beam-on AP positions over the entire delivery. (c,d) High-pass and low-pass filtering of the measured AP displacement curve used to separate rapidly varying motion components from slower baseline drift. (e,f) Probability distributions of the high-pass- and low-pass-filtered AP coordinates for all measured positions and beam-on positions. This representative comparison of measured and beam-on distributions illustrates how delivery sampling can modify the effective motion distribution used for dose reconstruction.

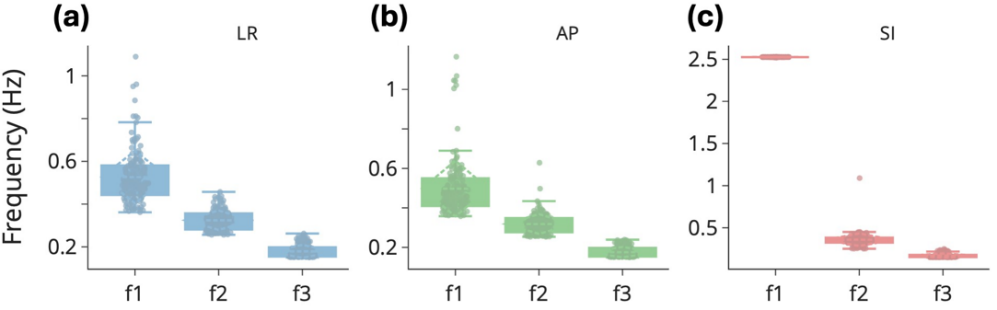

Figure S2. Dominant frequency components of intrafraction target motion across 150 fractions, 30 patients. The three largest spectral peaks identified from power spectrum analysis of the time-dependent displacement curves are shown for motion in the (a) left–right (LR), (b) anterior–posterior (AP), and (c) superior–inferior (SI) directions. Individual points represent fractions, and boxplots summarize the cohort distribution for the first, second, and third ranked frequency components (f1, f2, and f3). The highest-ranked SI frequency component was consistently observed at 2.5 Hz and was interpreted as measurement noise, whereas the remaining dominant frequency components were consistent with various modes of respiratory-related motion.

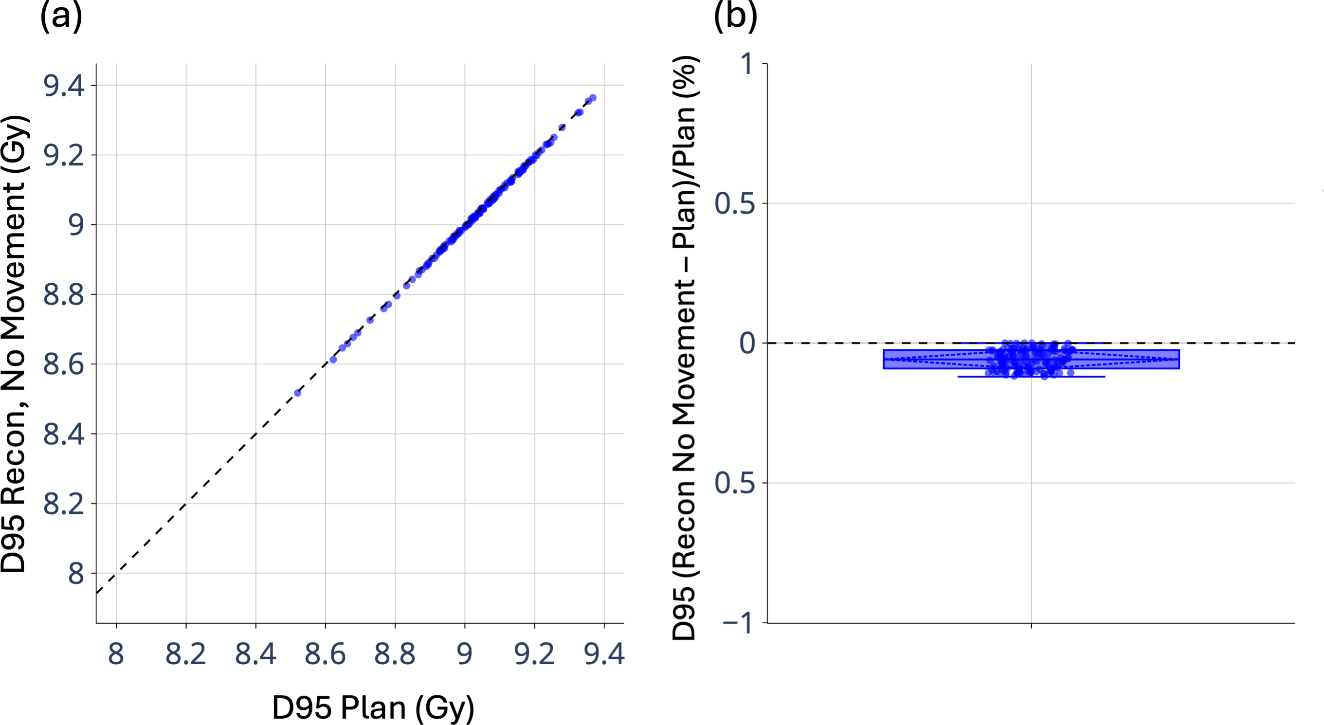

Figure S3. Validation of the dose-reconstruction pipeline under zero-displacement conditions. (a) Reconstructed D95 without motion plotted against planned D95 for all evaluated target instances; the dashed line denotes identity. (b) Distribution of normalized D95 differences, calculated as ΔD95 = (Recon − Plan)/Plan × 100%, with individual observations overlaid on the boxplot. Zero-displacement reconstructions closely reproduced the planned D95, with deviations limited to approximately −0.1% to 0%, indicating negligible numerical bias.

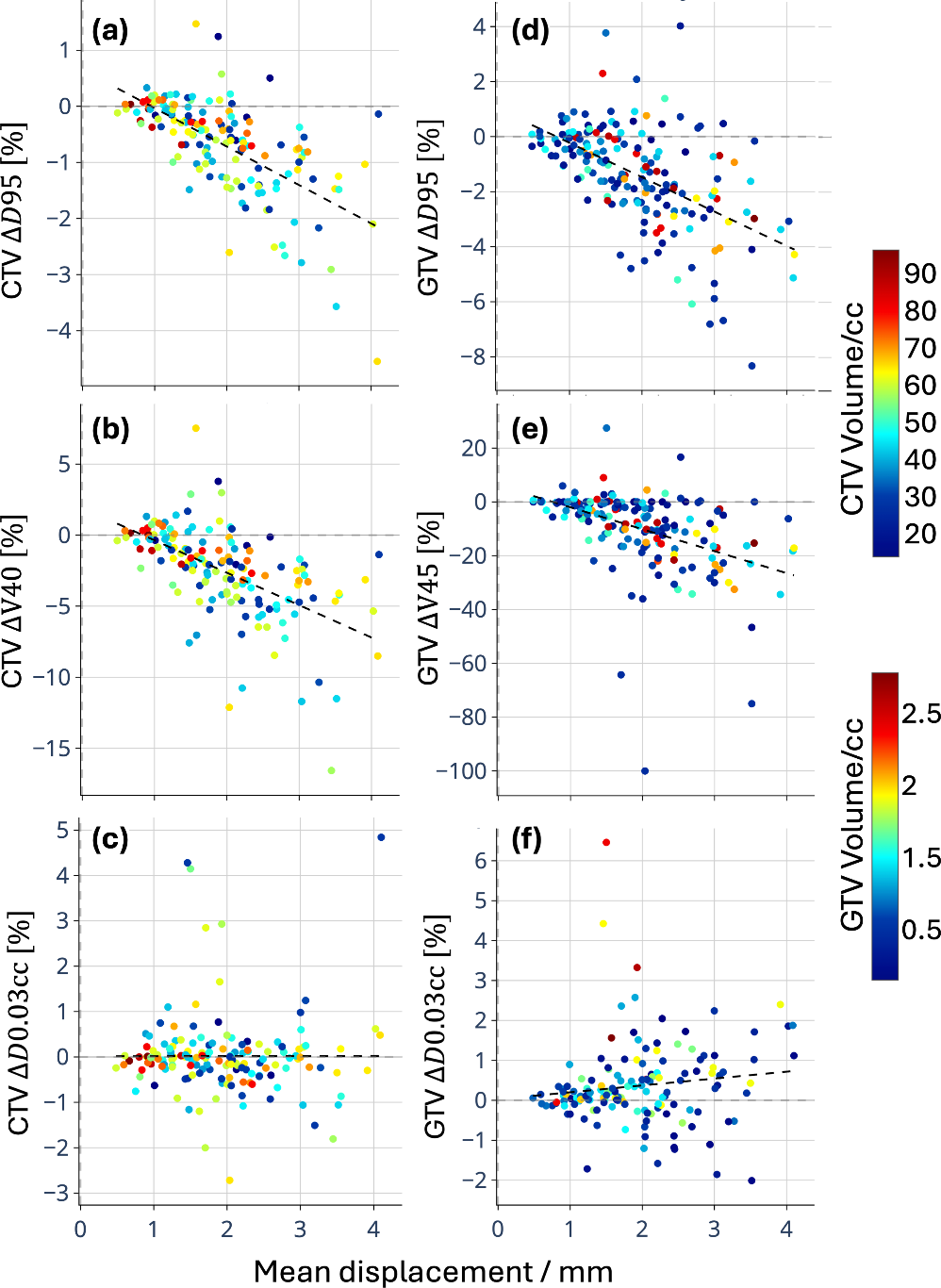

Figure S4. Correlation between mean intrafraction displacement and reconstructed dose deviations. Per-fraction dosimetric deviations, calculated as ∆Metric = (Recon − Plan)/Plan×100%, are shown as a function of mean target displacement for CTV metrics (a–c) and GTV metrics (d–f). Panels show changes in D95, prescription-dose volume coverage, and D0.03cc for the CTV (∆D95, ∆V40Gy, ∆D0.03cc) and GTV (∆D95, ∆V45Gy, ∆D0.03cc). Individual points represent fractions and are color-coded by structure volume; dashed lines denote linear mixed-effects regression fits. Reductions in target coverage metrics were significantly correlated (p<0.05) with mean displacement, whereas the correlation of ∆D0.03cc with mean displacement was not significant. The correlation between coverage metrics and structure volume was not significant; structure volume was a significant positive predictor of ∆D0.03cc for GTV, but not CTV. Negative values indicate reduced reconstructed dose relative to the planned dose.

### Patient- and physician-reported GU/GI toxicity events

Exploratory GU/GI toxicity events were summarized retrospectively. International Prostate Symptom Score (IPSS) values before and after treatment were compared for patients with available assessments. For each patient, the maximum post-treatment IPSS change was calculated as the maximum IPSS recorded after treatment minus the latest IPSS recorded before fraction 1. Genitourinary toxicity grades were extracted from the clinical record and summarized across the cohort. IPSS changes were grouped as >5, 1-5, 0, or <0 points and stratified by maximum genitourinary toxicity grade.

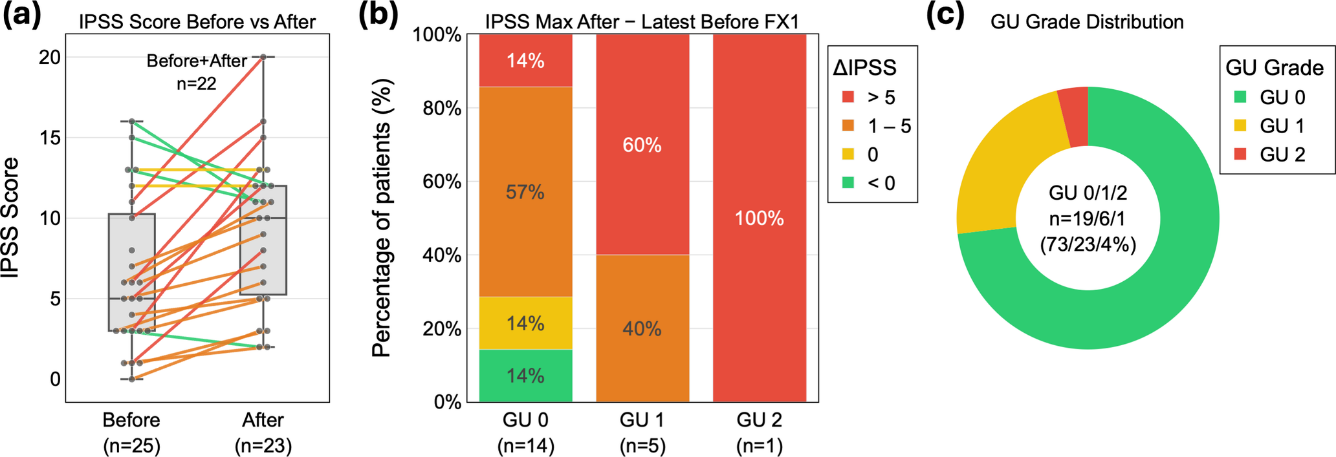

Figure S5. Patient-reported urinary symptom changes and genitourinary toxicity distribution. (a) International Prostate Symptom Score (IPSS) before and after treatment; box plots show the cohort distribution at each timepoint (Before, n=25; After, n=23) and connecting lines link patients with paired assessments (n=22), colored by IPSS change (∆IPSS: green <0, orange 1–5, red >5). (b) Distribution of the maximum post-treatment IPSS change — maximum IPSS after treatment minus the latest IPSS before fraction 1 — stratified by genitourinary (GU) toxicity grade (GU0, n=14; GU1, n=5; GU2, n=1) and grouped as >5, 1–5, 0, or <0 points. (c) Distribution of maximum GU toxicity grade across the cohort (GU0/1/2 = 19/6/1 patients; 73/23/4%). Sample sizes vary across panels because not all data were available for all patients.

Table S1. Summary of reconstructed dose deviations for target coverage metrics. Per-fraction and cumulative 5-fraction deviations are shown for GTV and CTV, calculated as ∆Metric = (Recon − Plan)/Plan. Values are reported as median with IQR in parentheses. Negative values indicate reduced reconstructed target coverage relative to the planned dose, with larger deviations observed for GTV coverage metrics than for CTV metrics. Cumulative deviations were smaller than per-fraction deviations, consistent with partial averaging of motion-induced dose differences over the treatment course.

|  |  | **Per-fraction** | **Cumulative (5 Fx)** |
| --- | --- | --- | --- |
| **GTV** | ∆V45Gy | -5.6% (-15.3 to -0.2%) | -4.0% (-13.8 to -0.5%) |
|  | ∆D95% | -1.2% (-2.4 to -0.1%) | -0.8% (-1.6 to -0.2%) |
| **CTV** | ∆V40Gy | -2.1% (-4.1 to -0.4%) | -1.2% (-2.9 to -0.7%) |
|  | ∆D95% | -0.5% (-1.1 to -0.1%) | -0.3% (-0.8 to -0.1%) |
